# Preparing correctional settings for the next pandemic: a modelling study of COVID-19 outbreaks in two high-income countries

**DOI:** 10.1101/2023.05.08.23289690

**Authors:** Jisoo A. Kwon, Neil A. Bretaña, Nadine Kronfli, Camille Dussault, Luke Grant, Jennifer Galouzis, Wendy Hoey, James Blogg, Andrew R. Lloyd, Richard T. Gray

**Author notes:** **Correspondence to:** CDr Jisoo Amy Kwon, Surveillance Evaluation and Research Program, The Kirby Institute, UNSW Sydney, Sydney NSW 2052, Australia.

## Abstract

Correctional facilities are high-priority settings for coordinated public health responses to the COVID-19 pandemic. These facilities are at high risk of disease transmission due to close contacts between people in prison and with the wider community. People in prison are also vulnerable to severe disease given their high burden of co-morbidities. We developed a mathematical model to evaluate the effect of various public health interventions, including vaccination, on the mitigation of COVID-19 outbreaks, applying it to prisons in Australia and Canada. We found that, in the absence of any intervention, an outbreak would occur and infect almost 100% of people in prison within 20 days of the index case. However, the rapid rollout of vaccines with other non-pharmaceutical interventions would almost eliminate the risk of an outbreak. Our study highlights that high vaccination coverage is required for variants with high transmission probability to completely mitigate the outbreak risk in prisons.

**Article Summary Line:** High vaccination coverage is required to eliminate the risk of an outbreak in prisons

## Introduction

Since the start of the COVID-19 pandemic in early 2020, correctional facilities around the world have experienced significant outbreaks of SARS-CoV-2 [1–5]. Such facilities (including gaols/jails, prisons, and other custodial settings), termed here “prisons”, are vulnerable to outbreaks of SARS-CoV-2 and other highly transmissible respiratory infections due to their congregate nature with unavoidable close contact between people. People in prison are particularly vulnerable to severe COVID-19 given the higher prevalence of co-morbidities and poorer social determinants of health compared to the general population [2, 6, 7]. Prisons’ enclosed environments mean that SARS-CoV-2 can easily spread between people in prison, correctional and healthcare staff (for an Australian prison setting and this terminology will be used throughout the manuscript)/correctional employees (in a Canadian prison setting), and visitors. The transfer of people in prison between correctional facilities and into the community can also fuel outbreaks in other facilities and into surrounding communities [8]. Prisons are therefore high-priority settings for coordinated public health responses to the COVID-19 pandemic and future outbreaks of other respiratory infections [3, 9–12]. However, the response to COVID-19 in prisons has been hampered due to limited access and use of personal protective equipment (PPE) in resource limited settings, poorer access and delay to the vaccination and vaccine hesitancy, security and logistical constraints, frequent movement of people between correctional settings, and the continuous entry and exit of people into the prison [13–17]. Correctional settings, therefore, require system-level and evidence-based responses [18, 19].

There have been several modelling studies evaluating the potential impact of prison-specific interventions to mitigate COVID-19 outbreaks in correctional settings. It was estimated that a large COVID-19 outbreak would be expected in prisons without both non-pharmaceutical interventions (NPIs) such as the use of PPE for people in prison and staff, decarceration of people in prison, quarantine at reception, isolation of people who are infected with COVID-19, and vaccination [20–24], particularly with delta and omicron variants [25–27], resulting in significant mortality [28]. These models, however, did not consider the heterogeneous transmission network inside prisons, the characteristics of the population (among whom there is an increased risk of severe disease), and, for the most part, failed to use real-world data for calibration. These previous studies also neglected to focus on the combination of public health interventions that could potentially mitigate COVID-19 outbreaks. To our knowledge, this is the first study which has sought to model a combination of intervention strategies using models validated with ‘real-world’ data. In this study, we aimed to develop a COVID-19 model for two high-income prison settings in Australia and Canada and validate the model outputs against outbreak data from these two settings, and sought to evaluate the potential impact of various intervention scenarios in averting cases and morbidity.

## Methods

We previously developed a COVID-19 Incarceration model by expanding on an existing spreadsheet model originally developed by Recidiviz (https://www.recidiviz.org) [29] to capture additional complex features within prison environments, to reflect the mixing patterns between people in prison and correctional and healthcare staff, and to model a broad range of interventions and mitigation strategies for COVID-19 outbreaks. The model is publicly available under an open-access license (GNU General Public License, Version 3) via GitHub [30] along with a user manual. This study was approved by the Human Research Ethics Committee at the University of New South Wales Sydney, Australia (HC200780). No additional approval from the McGill University Health Centre Research Ethics Board was necessary for the Canadian dataset as it was publicly available.

### Settings for the Australian and Canadian prisons

The Australian prison is a maximum-level quarantine prison with approximately 1,000 adult men (>18 years old). The Canadian prison is the largest provincial prison in Quebec, where it was the epicenter of the SARS-CoV-2 epidemic, with a capacity of 1,400 adult men (>18 years old) [31, 32]. Both prisons experienced SARS-CoV-2 outbreaks while multiple non pharmaceutical interventions (NPIs) were in place, but prior to a vaccine being available. A more detailed explanation of COVID-19 outbreaks in both prisons and interventions implemented will be explained later.

### Model structure

The scenarios and structures of the model were informed by a reference group drawn from both healthcare and correctional organizations. A detailed explanation of the model structure is available elsewhere [33]. Here we provide a summary and focus on the intervention scenarios investigated in the two settings. Briefly, the model is compartmental and implemented in Microsoft Excel (Redmond, WA). The model includes compartments representing the number of people in prison and staff who are susceptible, exposed, infectious, have mild illness, severe illness, are hospitalized, and recovered, with the number of deaths and new infections calculated daily (Figure 1). The model incorporates potential virus transmission between people in prison, correctional and healthcare staff, and visitors. It allows the designation of the prevalence of vulnerabilities in the population which could lead to severe COVID-19 disease, varied numbers of close contacts, and the daily intake and release of people in prison. People in prison are grouped by age in the model with a certain proportion in each age group considered ‘vulnerable’ to severe COVID-19 (classified as a patient of concern). Seven age-group cohorts were used for people in prison (0-19, 20-44, 45-54, 55-64, 65-74, 75-84, and 85+ years of age). The model incorporated the probability of showing symptoms, being hospitalized, and moving to critical or intensive care among those hospitalized (Table 1). Age-specific infection fatality rates were specified for each age group. As people in prison enter the prison, they are allocated to each age group based on the age distribution of the people currently incarcerated. The model is implemented in a difference equation framework with the number of people in each compartment updated daily over 120 days. The model tracks people in prison who enter the prison either through reception or via transfer from another correctional setting, the daily number of visitors, and correctional and healthcare staff working at the site. People in prison leave the model population to reflect the number that are released after the end of their sentence or released early as a public health mitigation measure. We assumed symptomatic people in prison were not released until recovered. Staff are assumed to attend the prison site every day. Individuals from the community can visit the site every day to represent family visitors, but they are assumed to only have a limited number of contacts each visit (with a family member in prison and correctional staff).

**Figure 1:**
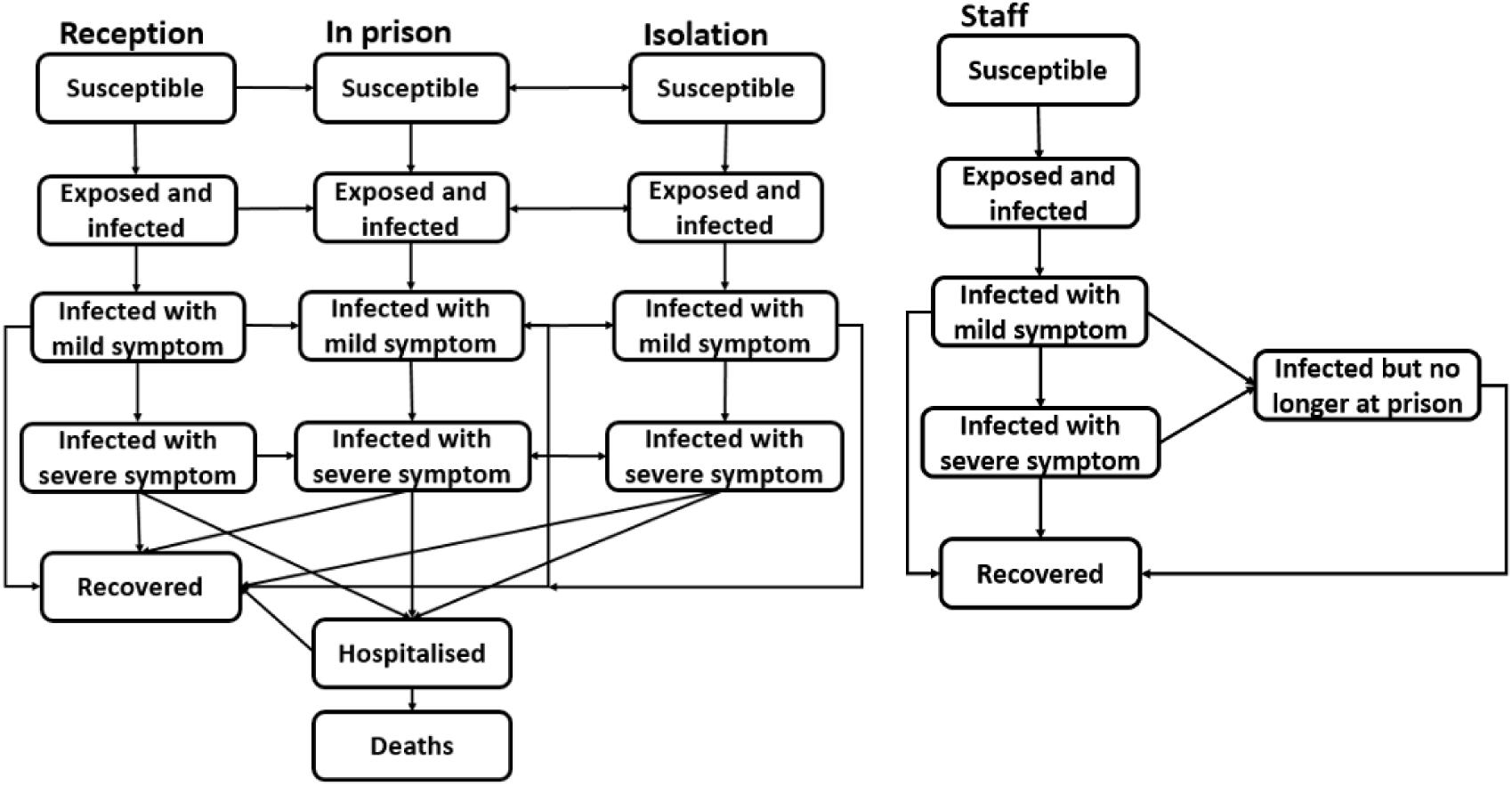
Schematic diagram of COVID-19 disease progression among people in prison and staff

**Table 1:**
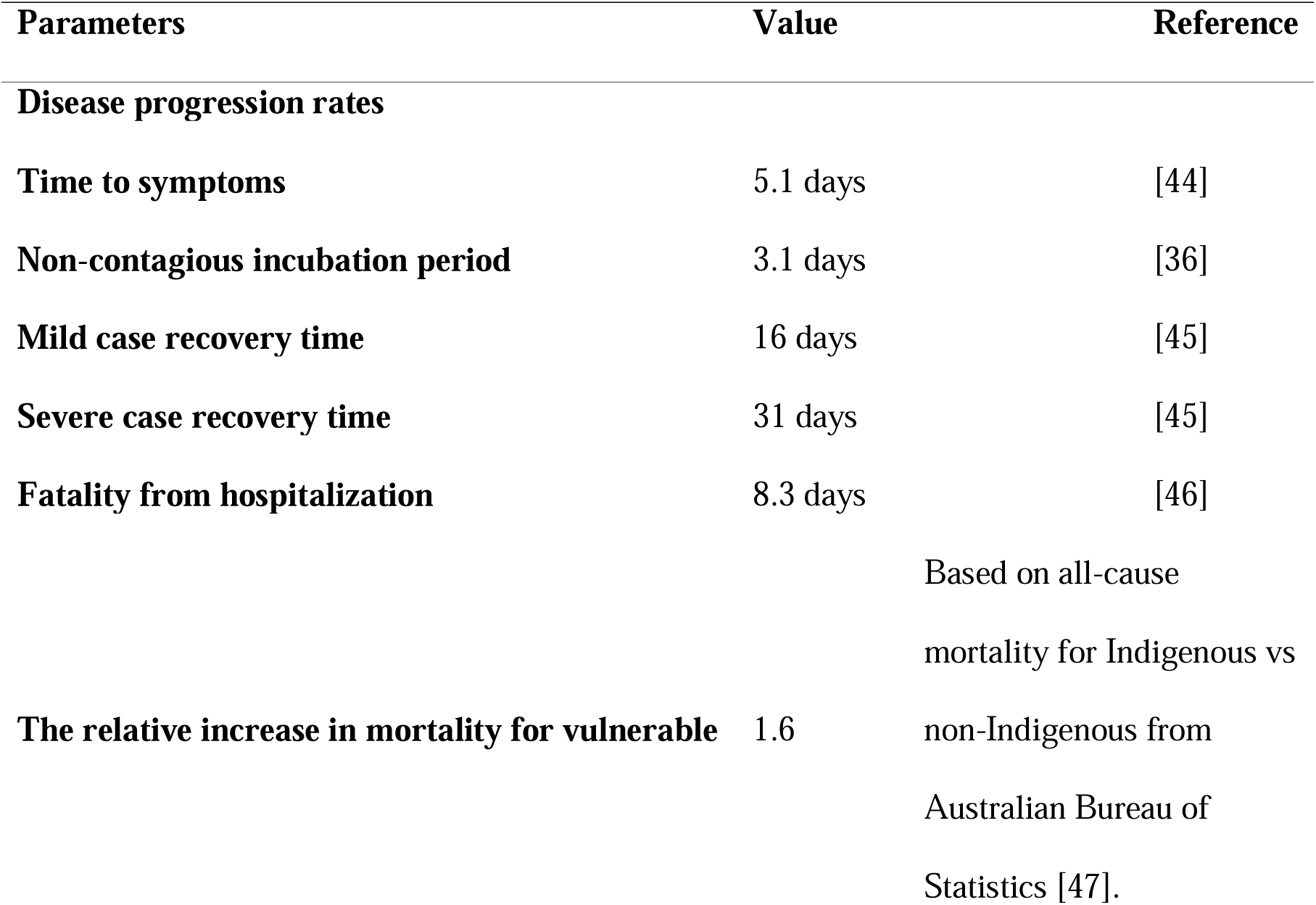
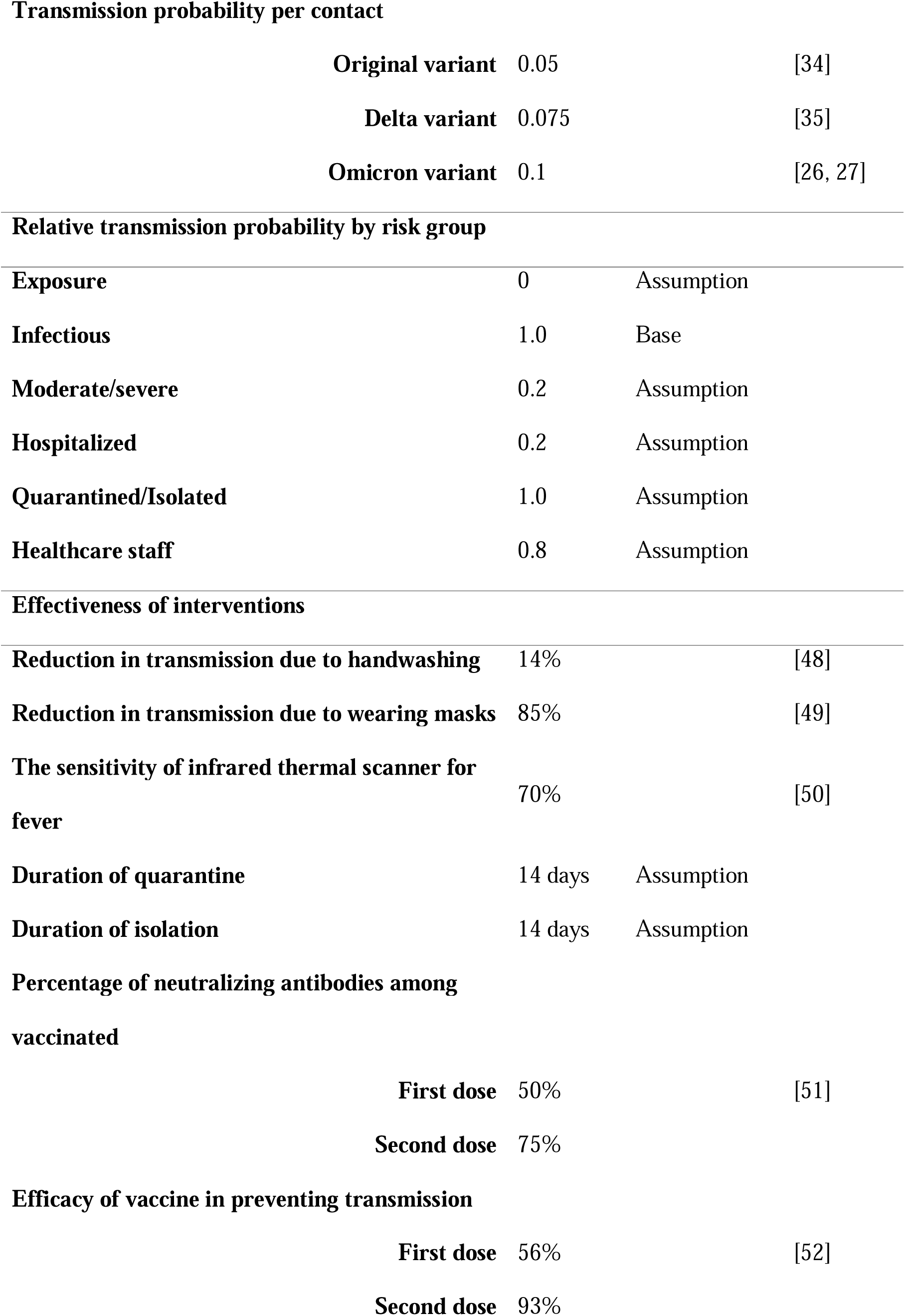
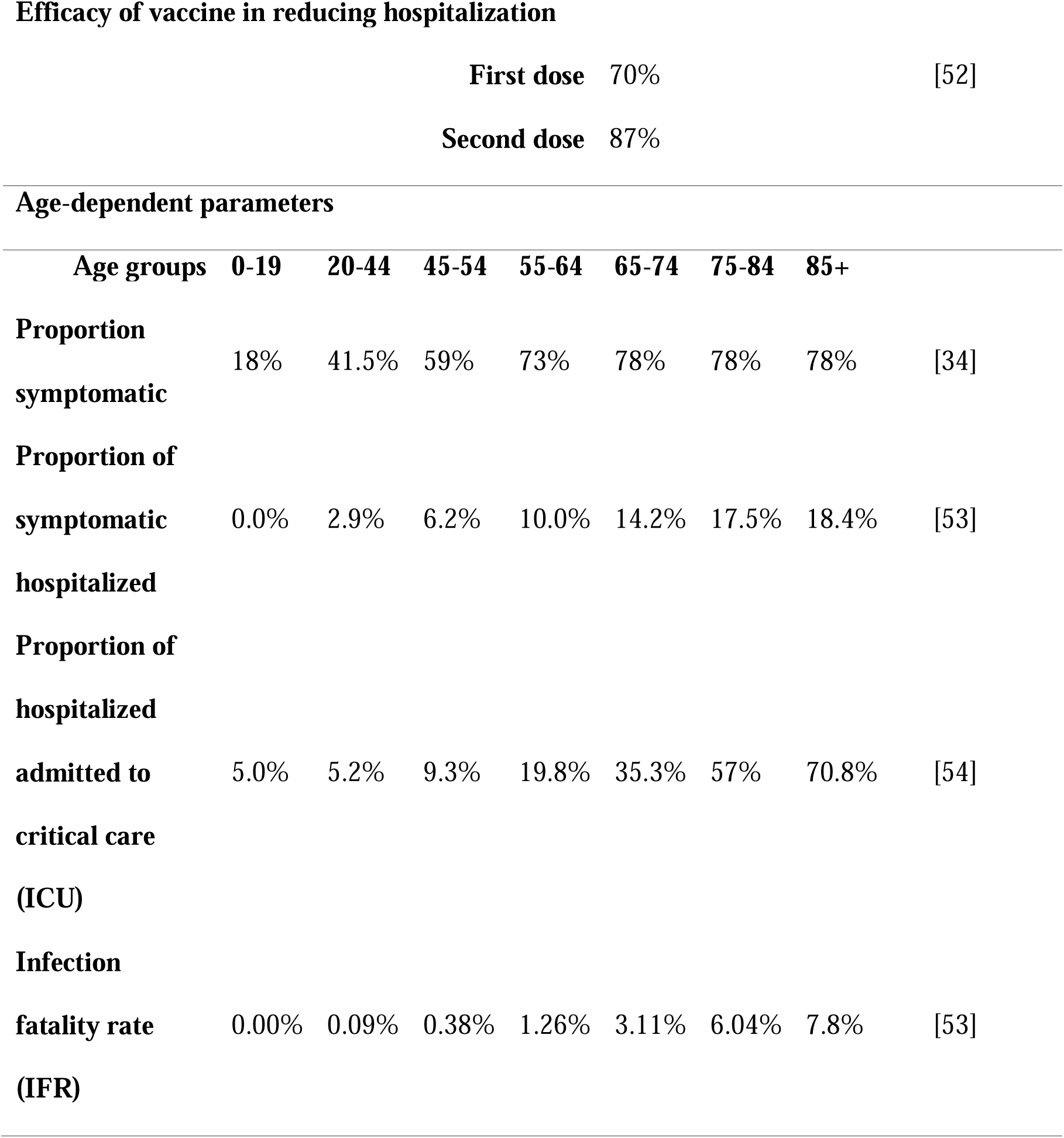
Model inputs and parameter estimates.

The COVID-19 progression rates were based on published data. The transmission of COVID-19 from infected to susceptible people per close contact with an infectious person had a value of 0.05 for the alpha variant [34], a value based on epidemics in Wuhan, China, accounting for different contacts through school, home, work and other contacts. While distance used for a close contact varies internationally, for the purposes of our analyses, we defined a close contact to be a distance of less than 1.5 meters for longer than 15 minutes. We assumed the transmission probability was 1.5 times higher for the delta variant [35], and two times higher for the omicron variant, compared to the alpha variant [26, 27]. This transmission probability was adjusted to reflect the variable of susceptibility by age, the use of PPE (including masks, hand washing, and personal hygiene measures) and disease stage (Table 1). Viral shedding during the course of infection was considered to be low during the exposed stage [36], and hospitalized patients (assumed to be isolated) and healthcare workers were assumed to always wear PPE (assumed to be 1 in the ‘Infectious’ stage and from 0 ‘Exposure’ to 0.8 among healthcare staff (Table 1)). The number of contacts is specified in the model for each population group, and we assumed homogeneous mixing within the modelled prison setting. The effect of vaccination in preventing transmission and reducing hospitalization among people in prison and staff receiving the first and second dose is detailed in Table 1.

### Interventions incorporated into the model

The effects of five intervention strategies to reduce the risk of COVID-19 transmission were incorporated into the model (Figure 2). All NPIs are delineated in light pink and vaccination in dark orange. These include: 1. Deferred incarceration or early release of people in prison (decarceration), 2. Use of PPE by staff or people in prison, including gloves and masks, 3. Quarantine of new people in prison at reception (assumed quarantine for 14 days for all newly admitted people in prison in single cells (preferred) or in groups (if quarantine capacity is limited), 4. Isolation of people in prison with suspected or proven infection (assumed isolation for 14 days), and 5. Vaccination of people in prison and staff. Each intervention was simulated individually or in combination (combining interventions from 1 to 4) for NPIs, and a combination of NPI with vaccination (combining interventions from 1 to 5).

**Figure 2:**
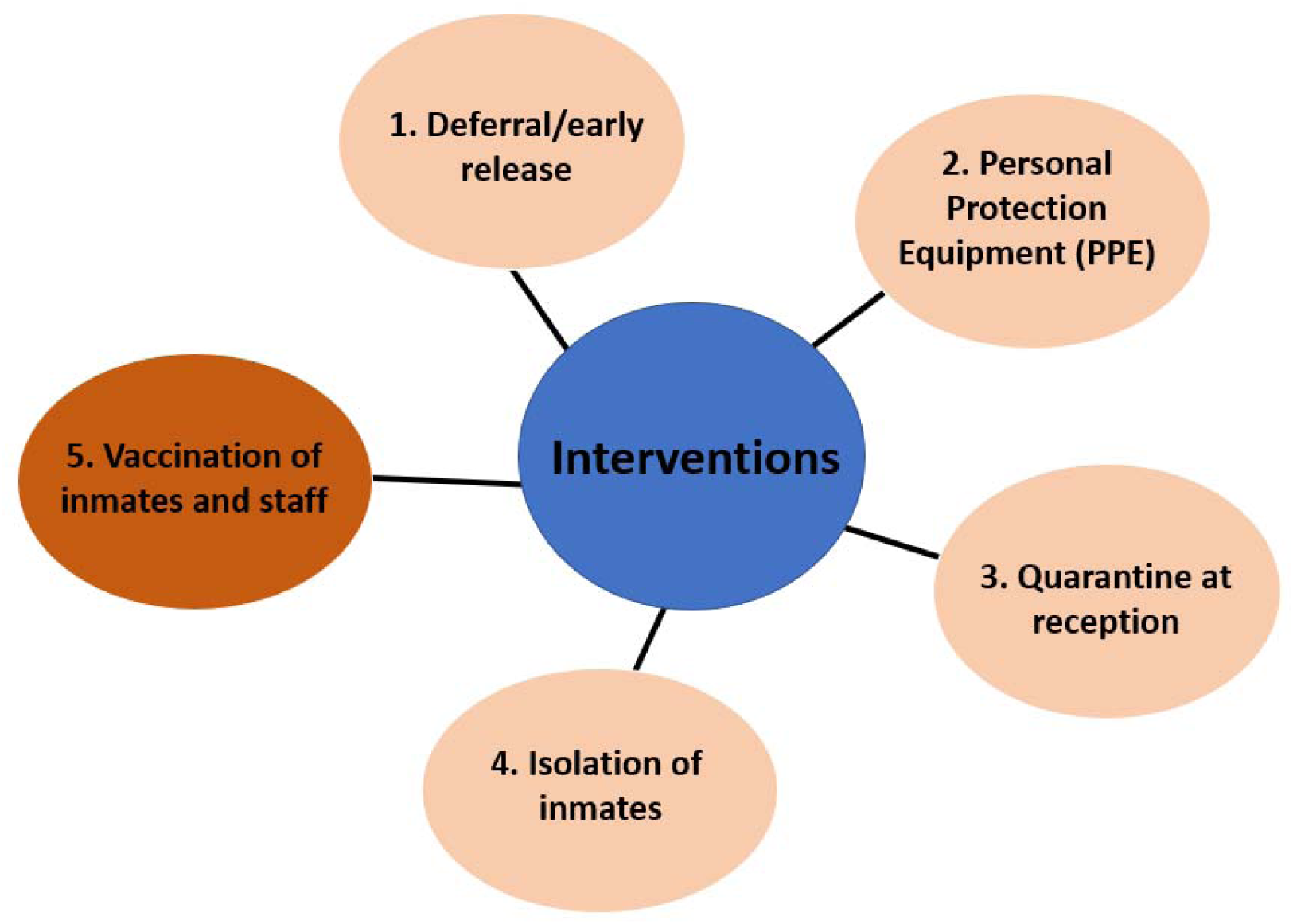
Interventions incorporated into the model. Each intervention and combined interventions were compared to the no response scenario (status quo). Non-pharmaceutical interventions are in light pink (NPIs) and vaccination is in dark orange.

### Demographic data

We collected data regarding demographics and prison characteristics from Corrective Services NSW (CSNSW) and the Justice Health & Forensic Mental Health Network (JHFMHN) for the Australian prison and the Ministry of Public Security [37, 38] for the Canadian prison. The number of contacts per person each day was estimated by the average number of people in prison in each cell, yard capacity; the size of work, training, or exercise groups; the number of patients each healthcare staff saw each day; the number of correctional staff working each shift and attending change-over meetings; and by surveys of staff and people in NSW prisons (collected separately and provided by CSNSW). Although staff work in shifts, they were assumed to intermingle extensively during each shift resulting in a high number of contacts and sufficient enough to transmit the virus during shifts. The number of contacts per healthcare staff) was estimated from the number of patients seen per day. It was assumed that 70-80% of healthcare staff have close contact with other correctional staff (personal communication with CSNSW and JHFMHN reference group). As this information was not available for the Canadian prison, we used similar intermingling and number of contacts per inmate and staff as the Australian prison as both facilities have similar structures and resources. We gathered all the detailed data explained above through consultation with the reference group which was then incorporated into the parameters.

### The Baseline scenario

The Australian prison experienced an outbreak with the delta variant from 11 August 2021 following multiple entries of infected inmates and staff. There were multiple NPIs in place at the time of the outbreak including: decarceration of people in prison, reduction in contacts, quarantine for 14 days at reception (entry), isolation of people in prison with suspected or proven infection, PPE for people in prison and staff, and thermal screening of non-essential staff and family visitors. Note that decarceration of people in prison in the Australian prison strategy was existed but the population size was not changed during the outbreak of COVID-19.

The Canadian prison experienced an outbreak during the early stages of the pandemic from 15 April 2020 when staff infected with the alpha variant entered the prison. Prior to this outbreak, there were several NPIs already in place aimed at controlling the number of close contacts each day, including: isolation among people in prison with suspected or proven infection, cessation of all visitors, 14-day quarantine of newly incarcerated people, and the distribution of PPE for all staff. Distribution of PPE to all people in prison was introduced in this prison during the outbreak from 2 June 2020 onwards.

To correspond to what is believed to have occurred and set a ‘baseline scenario’ for both the Australian and Canadian prison models (Figure 3), we used the prison-specific demographic data as well as the interventions in place at the time of each prison’s first outbreak. No vaccines were available in either prison at the time of the outbreak, however, vaccination began in the Australian prison among people in prison and staff during the outbreak and it likely contributed to mitigating the outbreak. In the Canadian prison, the first vaccine was administered on April 30, 2021 (personal communication on 19 January 2022, CIUSSS du Nord-de-l’Île-de-Montréal). A counterfactual ‘no-response’ scenario was run to see how large the COVID-19 outbreak could have been with no interventions in place (Figure 3).

**Figure 3:**
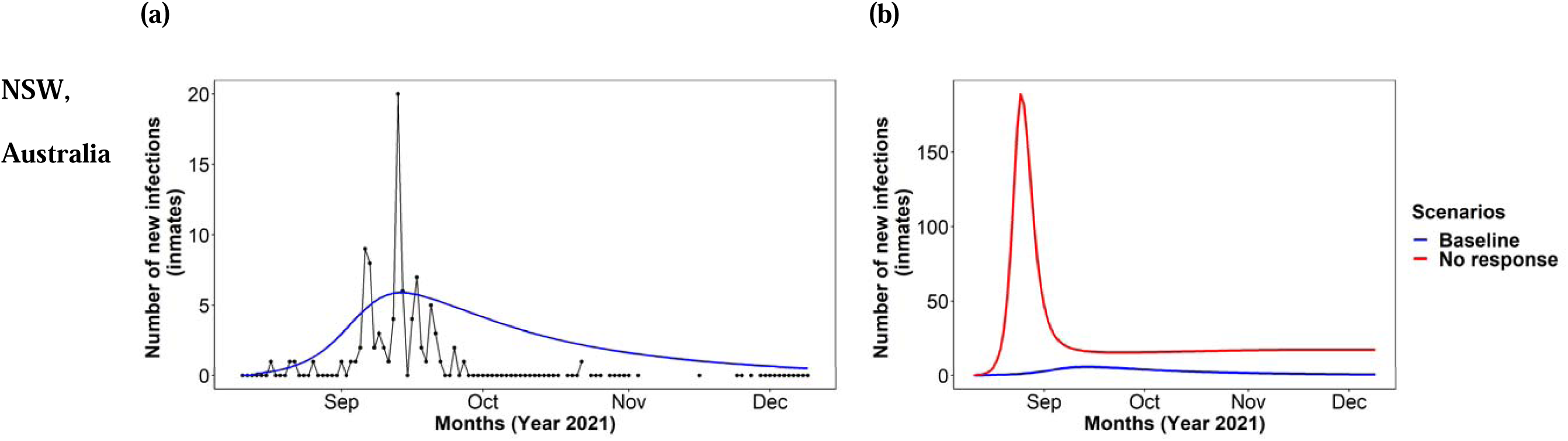
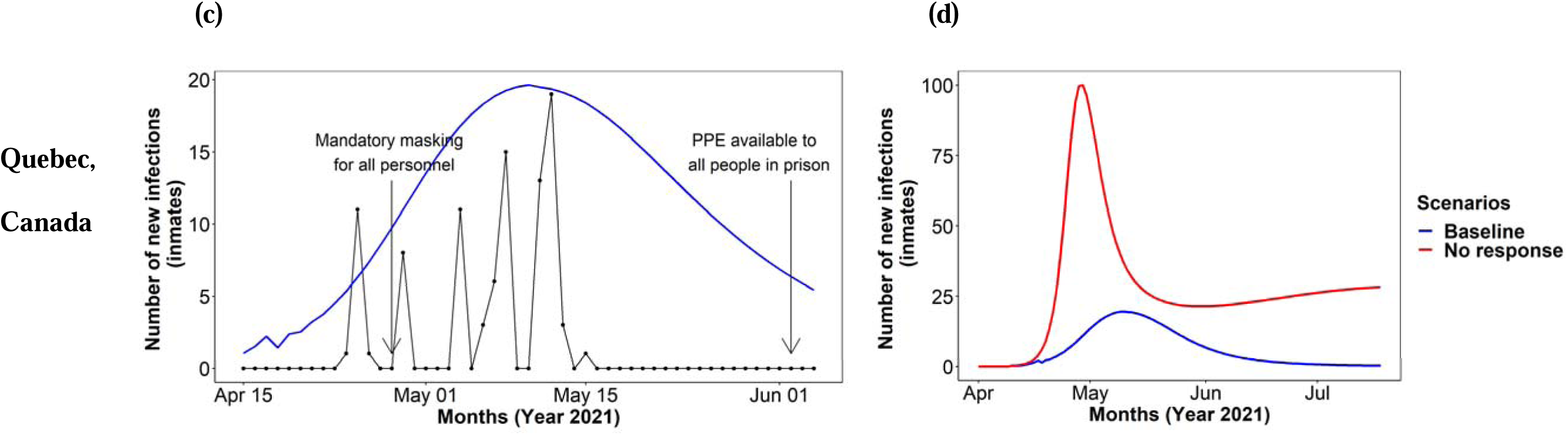
Number of new infections of COVID-19 among people in prison with the existing prevention strategies at the time of the outbreak (baseline, (a) and (c)) and comparison with the no intervention strategies (no response scenario (b) and (d)) in NSW, Australia andQuebec, Canada prisons. The baseline scenario in NSW, Australia was estimated with the delta variant and Quebec, Canada was estimated with the original variant.

### Applications of the model

#### Scenarios simulated

We simulated each model intervention separately (using scenarios 1 to 5) and in a combination scenario for 120 days to project the potential epidemic of COVID-19 within people in prison and staff. The number of hospital and intensive care unit (ICU) beds required are estimated from the model.

For the vaccination scenarios, on the advice of the reference group for the Australian prison, we assumed 50% of people in prison and 100% of staff were vaccinated (an estimate of the likely achievable coverage as vaccination of staff was mandated in the prison system). We assumed the same vaccination coverage among people in prison and staff in the Canadian prison as this information was not available. For intervention scenarios, we used the beta variant for both Australian and Canadian prisons to determine the impact of intervention strategies in both prisons. We further simulated a vaccination scenario using the omicron variant in both prisons to assess the possible impact of vaccination status in reducing COVID-19 outbreaks for variants with a higher transmission probability (Appendix).

## Results

### Model calibration and validation: impact of model assumptions in the Australian and Canadian prisons

Our model matched both the Australian and Canadian COVID-19 outbreaks well (Figure 3). In the Australian prison, where all NPIs were in place before the outbreak, the infections peaked on day 23 (Figure 3) with the first death from COVID-19 on day 26. The model estimated that there would have been 850 cumulative infections over 120 days with 1.7% of cases hospitalized at the peak of the infection (Table 2). In the Canadian prison, the infections peaked on day 28, with the first death from COVID-19 on day 33. The model estimated that there would have been 910 cumulative infections over 120 days with 80 people hospitalized at the peak of the infection (Table 2). Although the modelled estimates were higher than the number of people who were diagnosed with COVID-19 in the Canadian prison, we believe that there were undiagnosed cases in the prison (personal communication on 28th July 2020, CIUSSS du Nord-de-l’Île-de-Montréal). Therefore, the estimated modelling outbreak (blue line) was used as the baseline to assess the impact of the interventions compared to the baseline scenario.

**Table 2:**
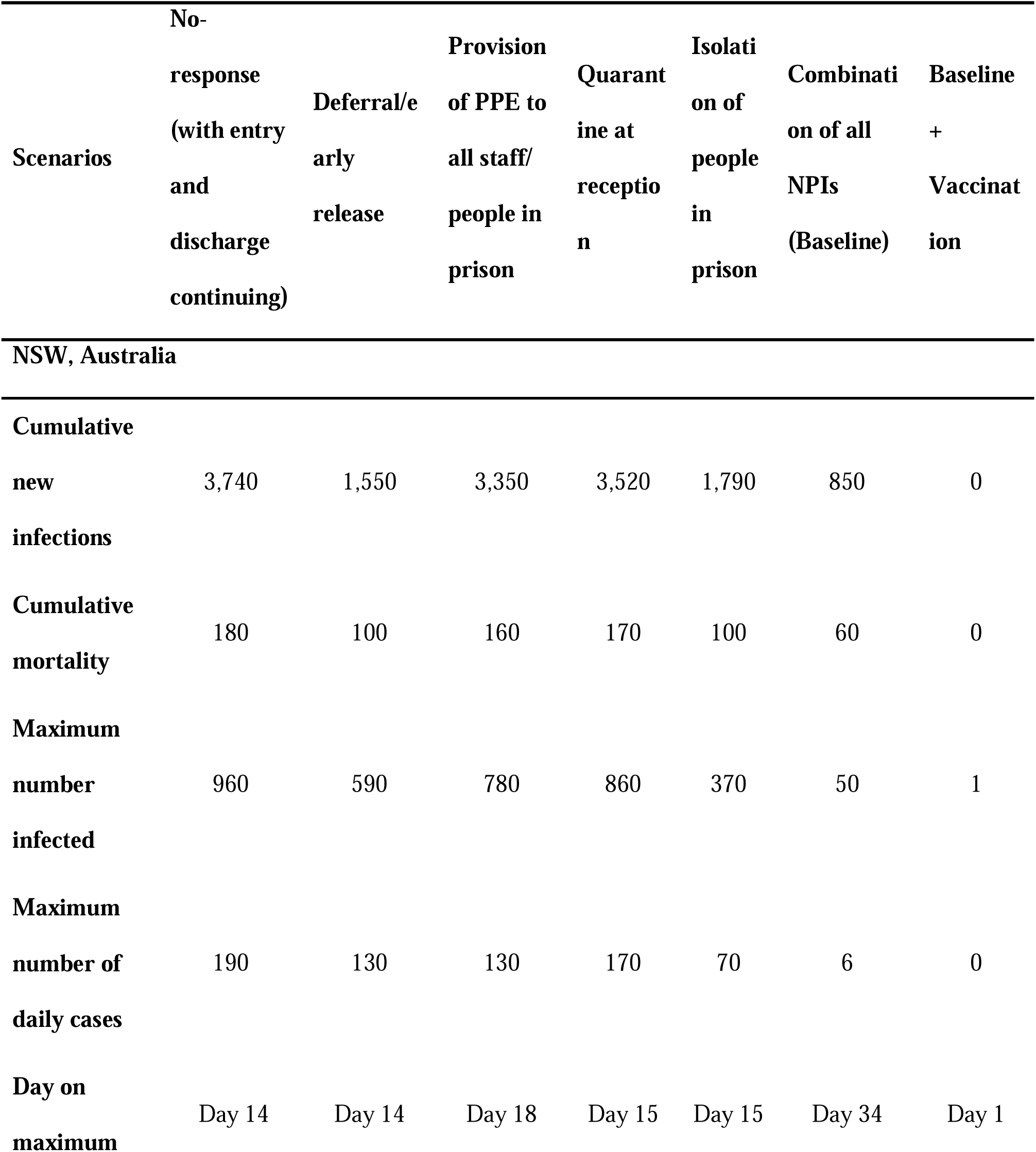
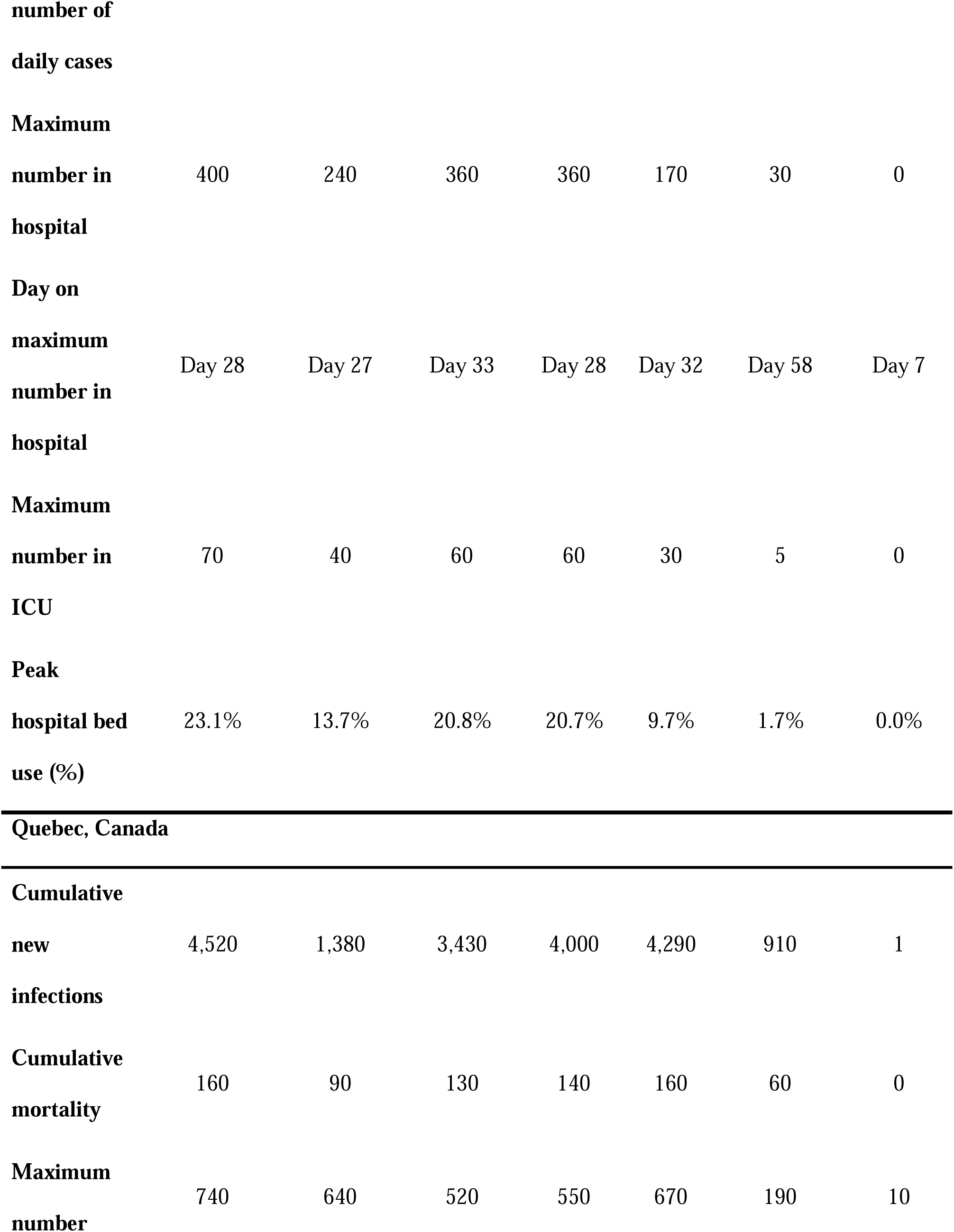
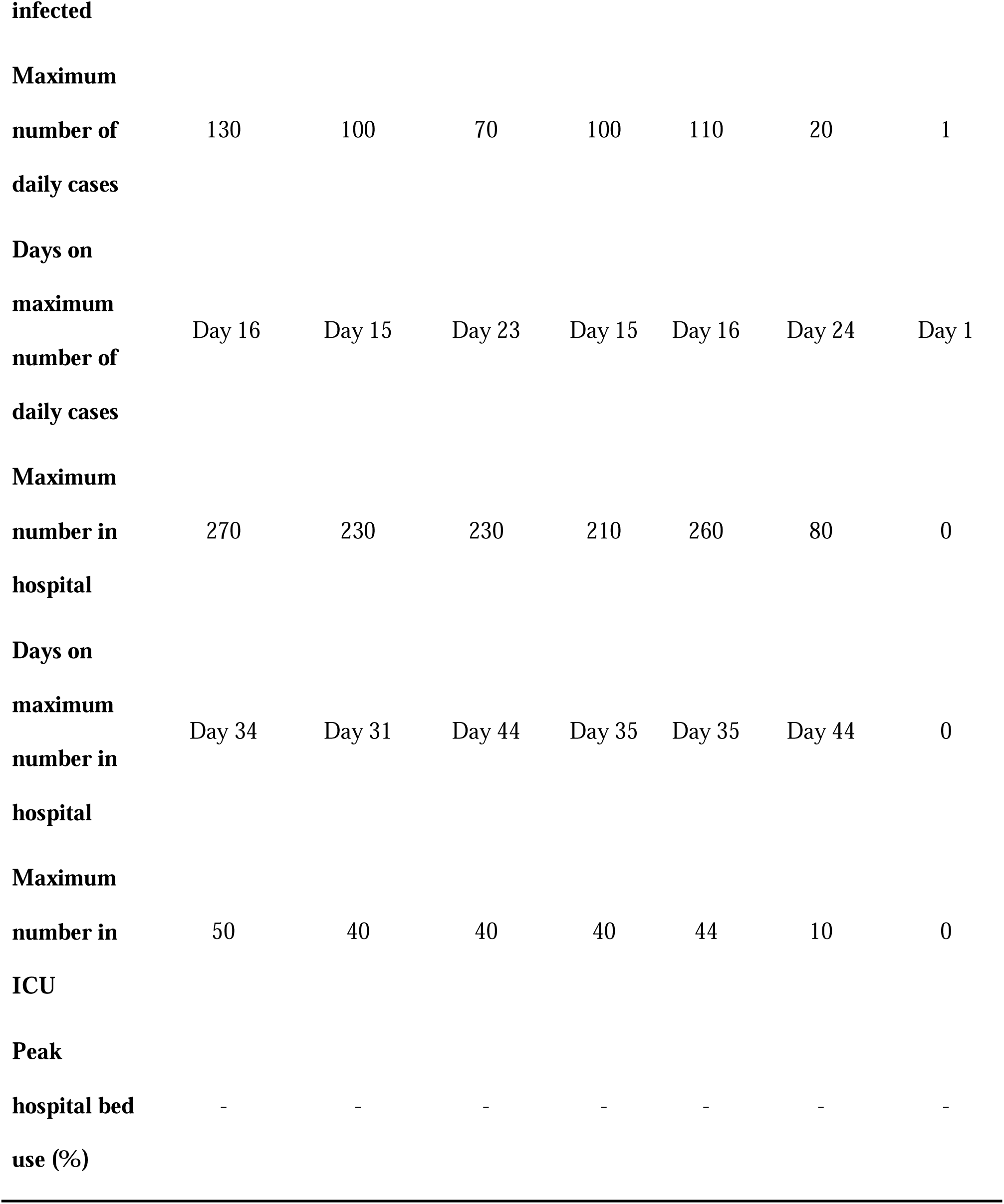
Key indicators from a COVID-19 outbreak (delta variant) in two prisons under the no-response and intervention scenarios (people in prison and staff). Each intervention is applied separately and in combination and run for 120 days. Results are for the overall population attending the prison site and are rounded to the nearest 10.

### Absence of interventions

#### Australian prison

Our model showed that, in the absence of any interventions (no response scenario, assuming admissions and releases continue), almost 100% of people in prison would become infected within 21 days of the outbreak, with 190 infections a day at the peak of the outbreak (day 14; Table 2 and Figure 3). Within 120 days of the outbreak, a total of 180 deaths due to COVID-19 were estimated (with deaths yet to plateau by 120 days; Table 2). At the peak of prevalence, approximately 84% of correctional and 92% of healthcare staff would also be infected and unable to attend work at the prison (Appendix, Figure A.1). Furthermore, our model estimated that if no response were in place, 470 hospital including 70 ICU beds would be needed at the peak of the outbreak in the local hospital facility (Table 2).

#### Canadian prison

For the following intervention scenarios in the Canadian prison, the delta variant was used to ensure consistency with the Australian prison. The model showed that, in the absence of a public health response (no response scenario), there would have been a large spike of COVID-19 cases (assuming admissions and releases continue) with almost 100% of people in prison becoming infected within 20 days (Figure 3). In this scenario, a total of 4,520 people in prison and staff would be infected during 120 days of an outbreak, with 160 deaths (Table 2). At the peak of the prevalence among people in prison, 94% of correctional and 83% of healthcare staff would also be infected and unable to attend work at the prison (Appendix A.1). Our model estimated that if no response were in place, 320 hospital including 50 ICU beds would be needed at the peak of the outbreak (Table 2).

### Interventions

#### Australian prison

The model predicted that reducing the prison population size (decarceration) had the biggest impact in reducing infections (among both people in prison and staff) in the Australian prison (59% reduction in cumulative incidence), followed by isolation of people in prison (52% reduction), PPE (11% reduction), and quarantine at reception (6% reduction in reception prison) (Figure 4 (a), Table 2). The model also showed that each intervention would reduce the number of occupied hospital and ICU beds (Table 2). In combination, the interventions (baseline) led to a substantially reduced outbreak with 77% fewer infections during the 120-day outbreak period compared to the no-response scenario (Figure 4 (a), Table 2).

**Figure 4:**
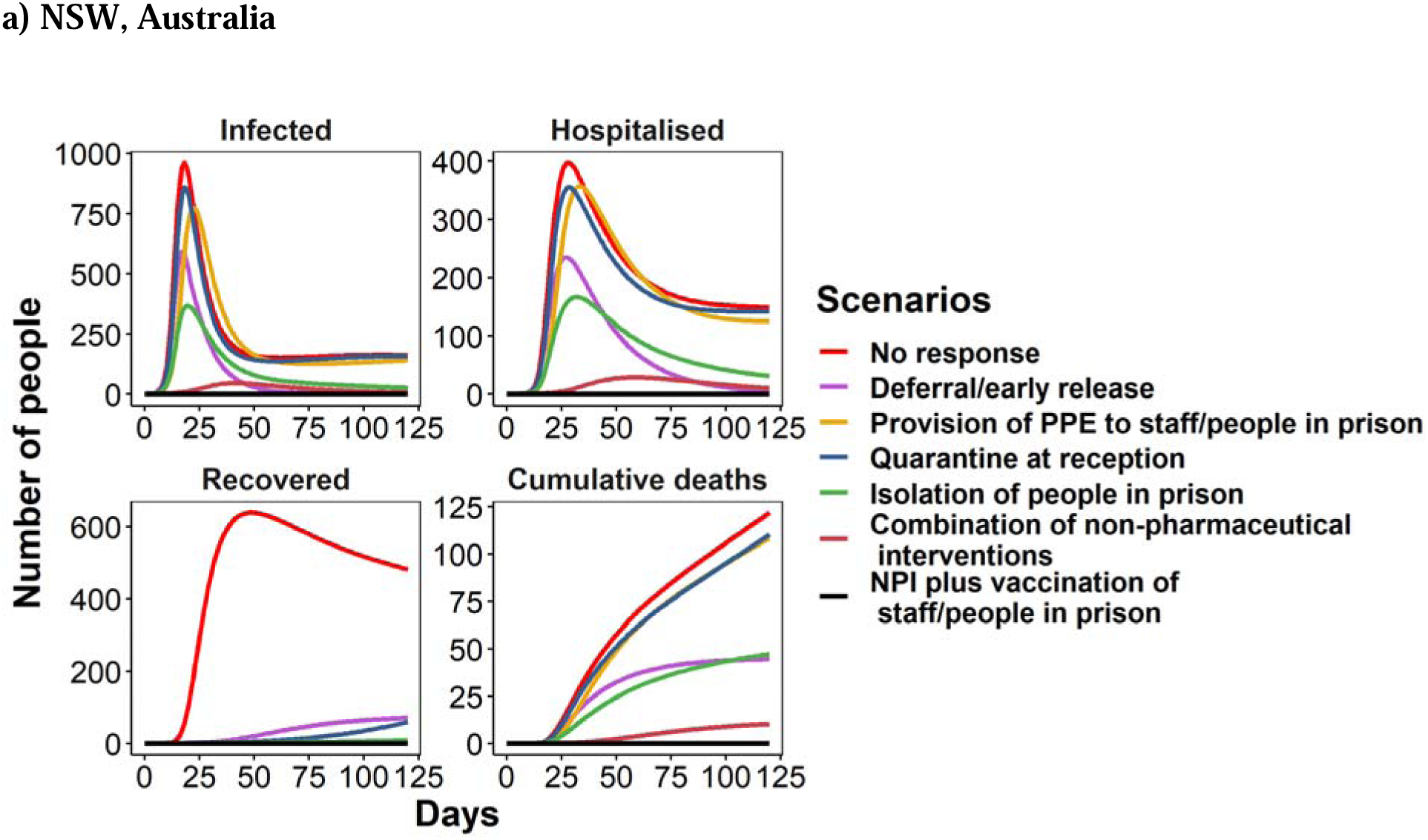
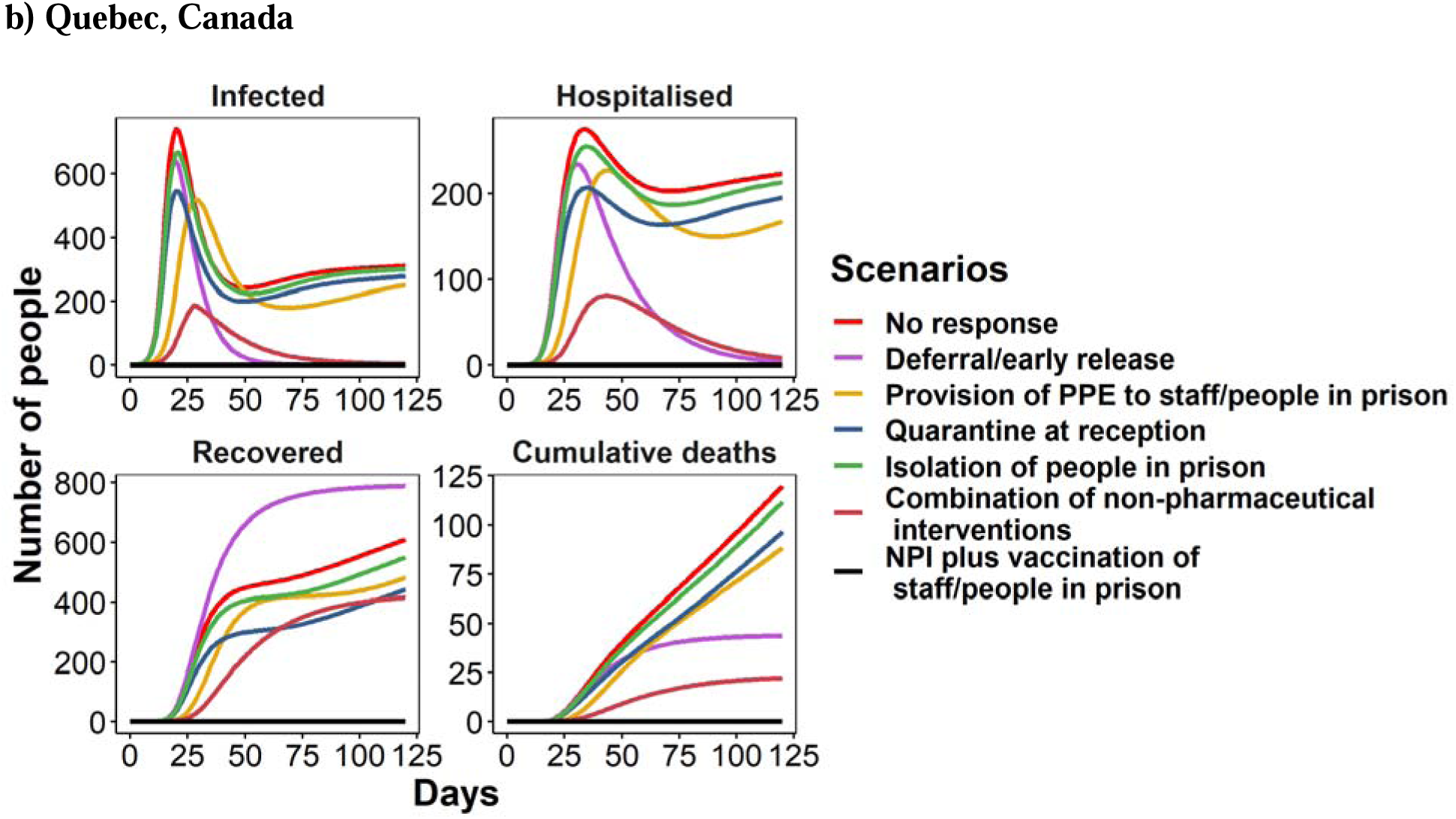
Change in the number of people in prison infected, hospitalized, recovered and the number who have died throughout a COVID-19 outbreak (delta variant) in the (a) NSW, Australia and (b) Quebec, Canada under the no-response and intervention scenarios. Each intervention is applied separately and in combination.

#### Canadian prison

In the Canadian prison, decarceration also had the biggest impact in reducing cumulative infection over 120 days (69% reduction), followed by PPE (17% reduction), quarantine at reception (12% reduction), and isolation of people in prison (5% reduction) (Figure 4 (b), Table 2). The impact of isolation was smaller in the Canadian prison than in the Australian prison due to the reduced capacity for isolation and quarantine of people in prison (a maximum of 100 inmates for isolation and 252 for quarantine in the Canadian prison compared to a maximum of 900 for isolation and quarantine in the Australian prison). In the baseline scenario, 80% of COVID-19 infections were averted compared to the no-response scenario over 120 days (Figure 4 (b), Table 2).

#### Vaccination coverage among people in prison with/without NPIs

The model predicted immunization of people in prison and staff would have a substantial impact on COVID-19 outbreaks. Ensuring at least 50% of people in prison are vaccinated with 100% of staff fully vaccinated (following a mandate which occurred in Australia but not in Canada) from day 0 in both prison settings would completely prevent an outbreak from occurring if other NPIs remain in place during the 120-day outbreak period (Figure 4, Table 2). Our model predicted that 50% vaccination coverage among people in prison is not enough to prevent COVID-19 outbreaks when transmission probability is high such as for the omicron variant, which has twice the transmission probability of the alpha variant (Appendix Figure A.2 & Figure A.3). Conversely, vaccinating 100% of people in prison and staff would prevent COIVD-19 outbreaks for both beta and omicron variants (Appendix Figure A.2 & Figure A.3).

#### Coverage of vaccination among staff

Ensuring 100% of correctional and healthcare staff are fully vaccinated (defined as a minimum of two doses of any mRNA vaccine) would mitigate transmission among staff even without other NPIs ensuring there would be minimal impact on the workforce (with delta variant, Appendix Figure A.4). In this scenario, almost 77% of the outbreaks among staff would be reduced but would still occur among people in prison, with a slower growth rate and a lower peak in daily infections, even if the outbreak was initiated by a staff member.

## Discussion

We developed a mathematical model incorporating the infrastructure of prison settings, COVID transmission, and disease dynamics. The model was used to assess combinations of targeted public health strategies to illustrate epidemic patterns and the effect of prevention or mitigation programs. The model can be readily adapted for application to different prison settings and to other respiratory viruses with similar transmission patterns, and so could be used for general pandemic preparedness in prison settings in the future. We applied the model to two prisons in two high-income countries - Australia and Canada, including the key characteristics of the prisons where real-world COVID-19 outbreaks occurred. Our model showed that modelling outputs predicted the COVID-19 caseload well, and highlighted the fact that there is a substantial risk of a major COVID-19 outbreak within prisons if an infected person/staff enters in the absence of control measures. Most importantly, our model demonstrated that NPI combined with vaccination (completely prevent an outbreak from occurring) was the most effective interventions, followed by decarceration (59-69% reduction in cumulative incidence over 120 days) in reducing COVID-19 outbreaks in prison settings.

The model showed that NPIs, and in particular, decarceration, can reduce the size of a COVID-19 outbreak within prisons and substantially reduce associated morbidity and mortality. Reducing the prison population size (decarceration), quarantine of people in prison at reception, and isolation of symptomatic people in prison are designed to reduce close contacts between infected and susceptible individuals—and essentially reduce the susceptible population within a prison, while the widespread use of PPE and vaccination reduces the risk of transmission during close contact. While these interventions are effective, our modelling showed that an outbreak could still occur. Our findings are important for resource-limited settings where access to vaccines in correctional settings may not available. For example, in Cambodia, prison authorities were urged to take action to reduce COVID-19 outbreaks [39] as the average prison occupancy was greater than three times its capacity. Hence, a decision to release people from prison (primarily those who pose minimal risk to public safety) was made to reduce overcrowding [39]. Reducing the number of people entering or leaving prisons to minimize the change of the COVID-19 outbreaks was also introduced in NSW prisons in the early outbreaks [40]

Where possible, vaccination of both people in prison and staff, combined with NPIs, will mitigate all future outbreaks and should be prioritized in countries where this has not occurred [41]. Our model highlighted that at the peak of prevalence among people in prison, 94% of correctional and 83% of healthcare staff would be infected and unable to attend work at prison. The loss of correctional and healthcare staff in person due to COVID-19, not only jeopardizes the safety and wellbeing of incarcerated individuals but also poses a significant threat to the public health of the wider community, as the virus can easily spread beyond the prison walls through staff members who may unknowingly carry the infection outside of the facility. Additionally, the absence of correctional staff can lead to a breakdown of order and security within the prison, making it more difficult to maintain the safety and rehabilitation of people in prison. Thus, it is crucial to prioritize the health and safety of correctional staff in order to ensure the overall well-being of both those incarcerated and the general public. It is also important to note that maintaining a high level of booster uptake is essential to ensure the immunity of both people in prison and staff in correctional settings which will help mitigate the risk of new outbreaks occurring.

A recent modelling study in the United Kingdom assessed the impact of vaccination, combined with various restriction levels (different rules in place to reduce close contacts including opening of non-essential shops, retails, traveling throughout the country or abroad, national lockdown), in reducing the number of people hospitalized and deaths due to COVID-19 [42]. The model highlighted that even with low level restrictions, vaccination can prevent number of people being hospitalized. The feasibility of achieving this comprehensive approach will likely vary across prisons, depending on the available resources, health and correctional infrastructure, and nature of operations within the facility. Another modelling study also showed that the combination of NPIs and vaccination can prevent deaths due to COVID-19, but required immense effort [20, 43]. For example, more than doubling of the vaccination rate was needed to halve the deaths within 100 days [43]. Therefore, it is evident that ongoing NPIs are needed in prison even if the vaccination rates are high, particularly with the emergence of increasingly transmissible COVID-19 variants.

There are some limitations to our study. It is important to note that as our model is compartmental, it does not capture all the complexities within a prison, or the specific interactions between individuals. This means it may overestimate the magnitude of an outbreak in a prison where the internal structure includes multiple wings and yards that can be isolated from each other in the event of an outbreak. Our model is also deterministic which means it does not capture probabilistic effects when the infection numbers are small. The model describes the movement between quarantine and isolation and the general prison population as an average rate equal to the inverse of the quarantine/isolation period. This means that there can be a slow release of infected individuals from quarantine/isolation in the model catalyzing an outbreak earlier than what might be expected. Depending on the intervention parameters, these are shown as a delayed trajectory with a slightly lower peak. However, people in prison in quarantine/isolation may still interact with staff, and exposed individuals may be released at the end of their quarantine/isolation periods, meaning that this slow spread of infection from quarantine/isolation is not unrealistic. Our model did not take into account the reduction in population size resulting from policing and court orders during COVID-19 outbreaks. For our next study, we plan to develop a more detailed individual-based model that considers the movement of inmates between prisons and courts. Finally, our model does not describe the impact of varied testing strategies for COVID-19.

Our study has several strengths. While our model was designed to investigate interventions for SARS-CoV-2 transmission in prisons, its structure is flexible enough to consider other respiratory infections in other closed population settings by changing the transmission probability. It also ensures flexibility to define and assess different scenarios and a combination of targeted public health strategies to illustrate epidemic patterns and the effect of prevention or mitigation programs. Here, we focused on COVID-19 outbreaks in two ‘real-world’ prison settings with intervention strategies to mitigate future outbreaks. Finally, the Australian and Canadian prisons were both male prisons, however, model inputs were based on published data for both females and males. Therefore, our findings are likely generalizable to female prisons.

In conclusion, our analysis demonstrates that the entry of one infected person into prison is sufficient to establish an outbreak, infecting almost all people in prison within 120 days in the absence of an intervention. A high vaccination coverage, in combination with other NPIs, would eliminate the risk of an outbreak in a prison, but the feasibility of these interventions will depend on both the health and custodial infrastructure of the facility. Lessons learnt from this study can be used to evaluate other respiratory viruses in congregate settings in the future.

## Data Availability

All data produced in the present work are contained in the manuscript

## Acknowledgements

JAK produced results, interpretation of findings, and wrote the manuscript; NAB provided interpretation of findings and manuscript writing; NK assisted in data collection, interpretation of findings, and manuscript writing; CD assisted in data collection, interpretation of findings, and manuscript writing; LG led the study, providing oversight in the design, implementation, interpretation of findings, and manuscript writing; JG, WH, and JB assisted in data collection and editing the manuscript; ARL led the study, providing oversight in the design, implementation, interpretation of findings, and manuscript writing; RTG led the study, providing oversight in the design, implementation, interpretation of findings, and manuscript writing. We acknowledge Nicola Archer-Faux from Department of Communities and Justice, Kimberley Conlan from Corrective Services NSW, Joshua Taylor from NSW health, Colette McGrath from Justice Health Forensic Mental Health Network NSW, Kevin Corcoran from Corrective Services NSW, Andrew Warren and Justine Kunz from the Recidiviz team, and David Boettiger from the Kirby Institute, UNSW for their input.

## Disclaimers

This study was funded by Corrective Services NSW. The Kirby Institute is funded by the Australian Government Department of Health and is affiliated with the Faculty of Medicine, UNSW Sydney. The views expressed in this publication do not necessarily represent the position of the Australian Government.

## Biographical Sketch

Dr. Amy Kwon is a mathematical modeler (lecturer) in Sydney. Her main research focuses on the development of mathematical models to understand the epidemiology of infectious disease and predict the impact and cost-effectiveness of interventions. She also leads other modelling projects to design intervention strategies and their impacts to assist governments in making informed decisions that affect the annual budget and resource allocation.

