## Supplementary Material for "Preparing correctional settings for the next pandemic: a modelling study of COVID-19 outbreaks in two high-income countries"

Jisoo A. Kwon^1^, Neil A. Bretaña^2^, Nadine Kronfli^3,4^, Camille Dussault^3^ Luke Grant^5^, Jennifer Galouzis^5^, Wendy Hoey^6^, James Blogg^6^, Andrew R. Lloyd^1^, Richard T. Gray^1^

1. Kirby Institute, UNSW Sydney, Sydney, New South Wales, Australia
2. Allied Health and Human Performance, University of South Australia, Australia
3. Centre for Outcomes Research and Evaluation, Research Institute of the McGill University Health Centre, Montreal, Quebec, Canada
4. Department of Medicine, Division of Infectious Diseases and Chronic Viral Illness Service, McGill University Health Centre, Montreal, Quebec, Canada
5. Corrective Services NSW, Australia
6. Justice Health Forensic Mental Health Network NSW, Australia

**Figure A.1: Number of staff available to work following the entry of one infected inmate (blue line: vaccinating 100% staff only without NPIs and red line: no vaccination for staff without NPIs; (a) NSW, Australia and (b) Quebec, Canada, if an outbreak was initiated by an inmate**

| (a) | 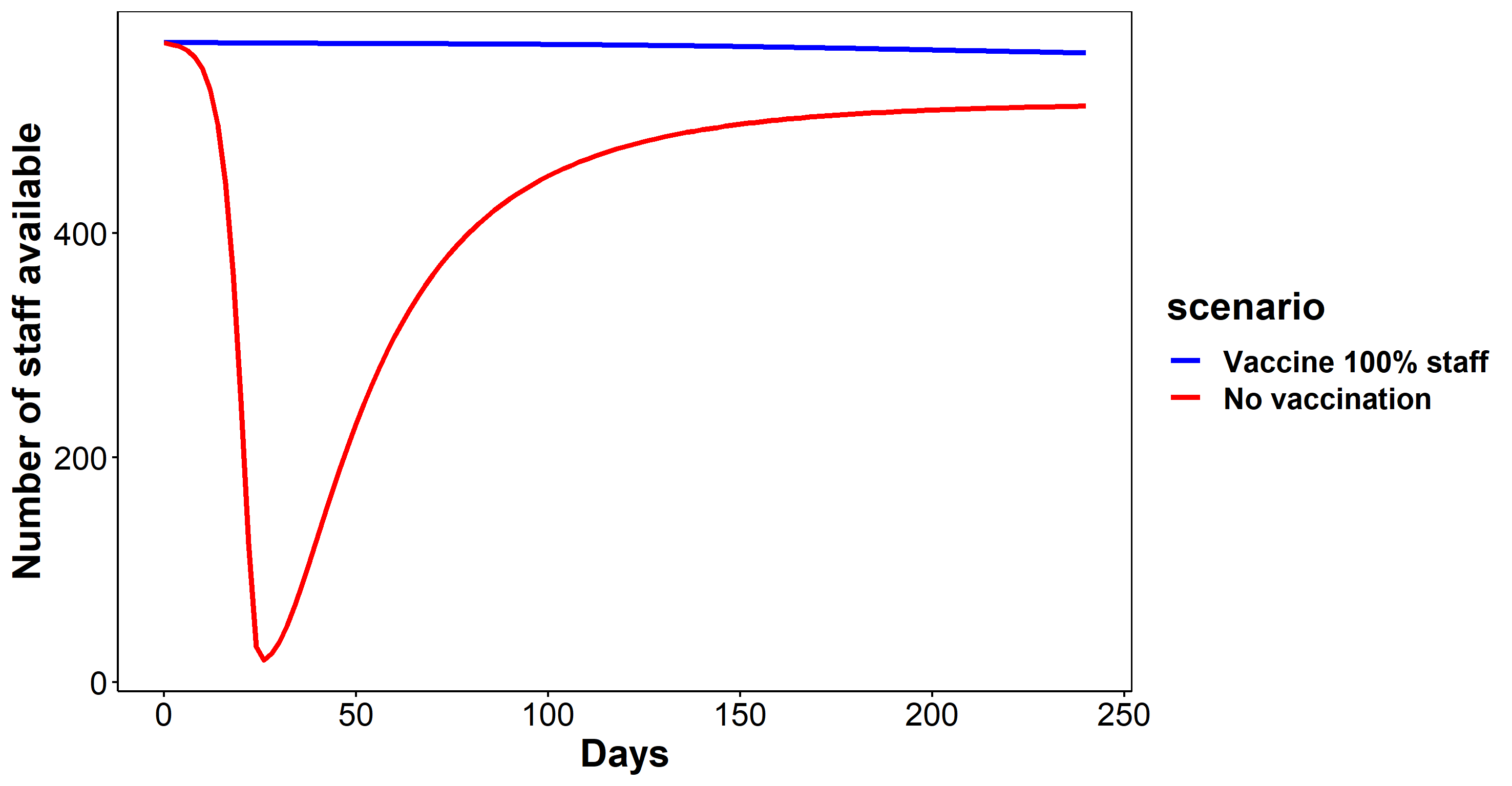 |
| --- | --- |
| (b) | 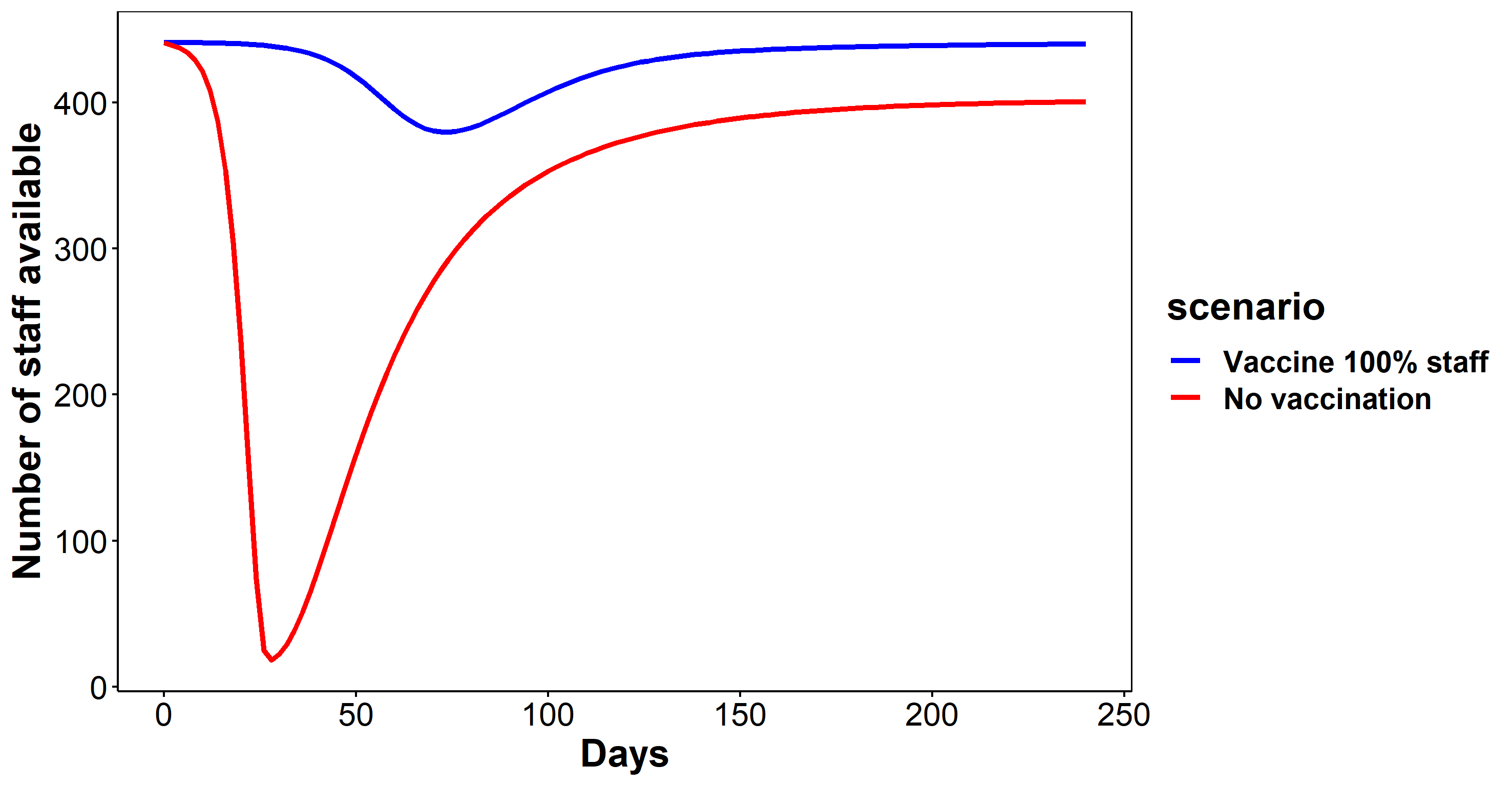 |

**Figure A.2: Number of infections (inmates, omicron variant) with vaccination scenarios following the entry of one infected inmate (red line: baseline line (50% vaccination of inmates) with NPIs, yellow line: 50% vaccination of inmates without NPIs, and blue line: 100% inmate vaccination of inmates without NPIs); (a) NSW, Australia and (b) Quebec, Canada**

| (a) | 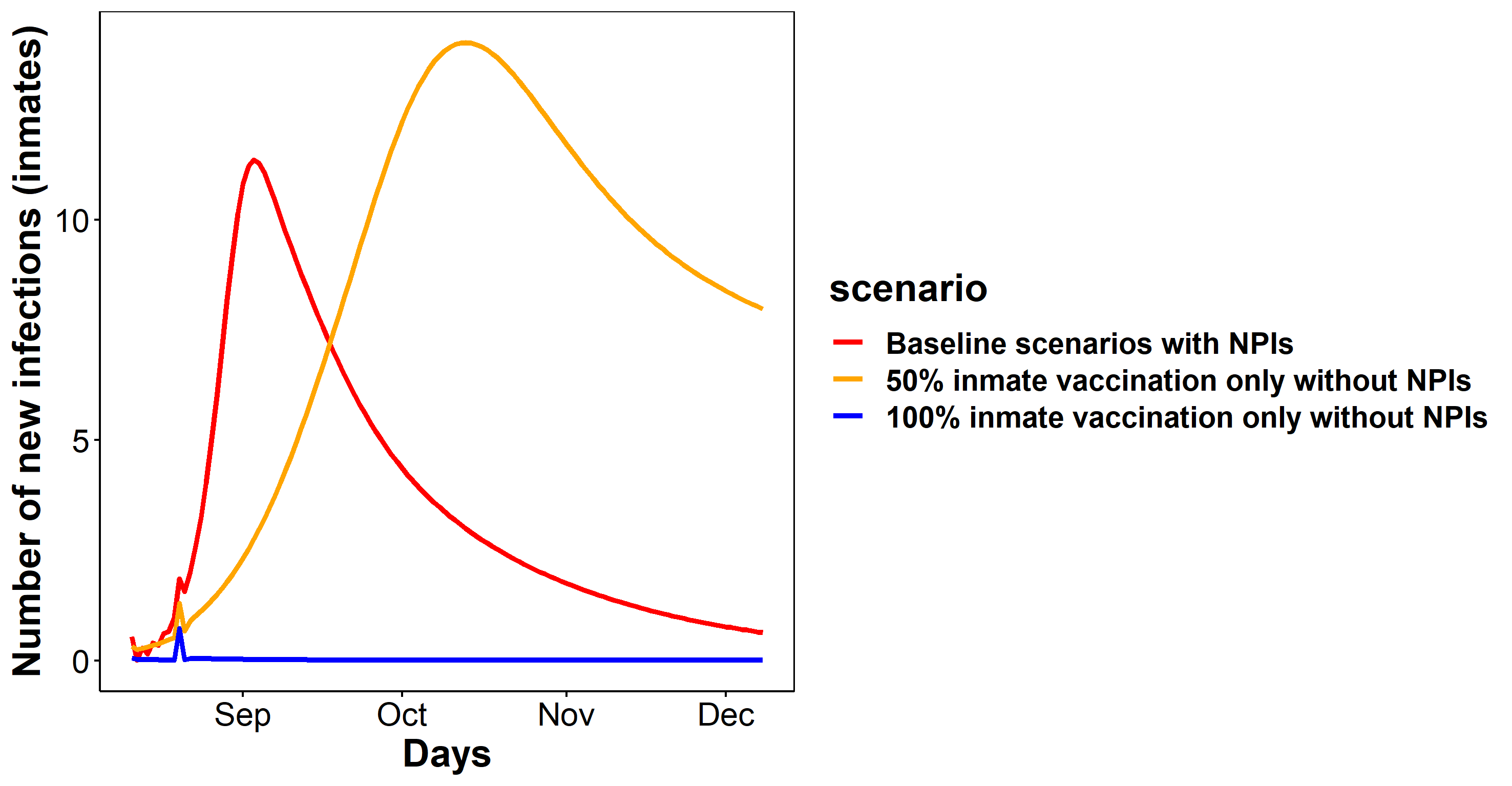 |
| --- | --- |
| (b) | 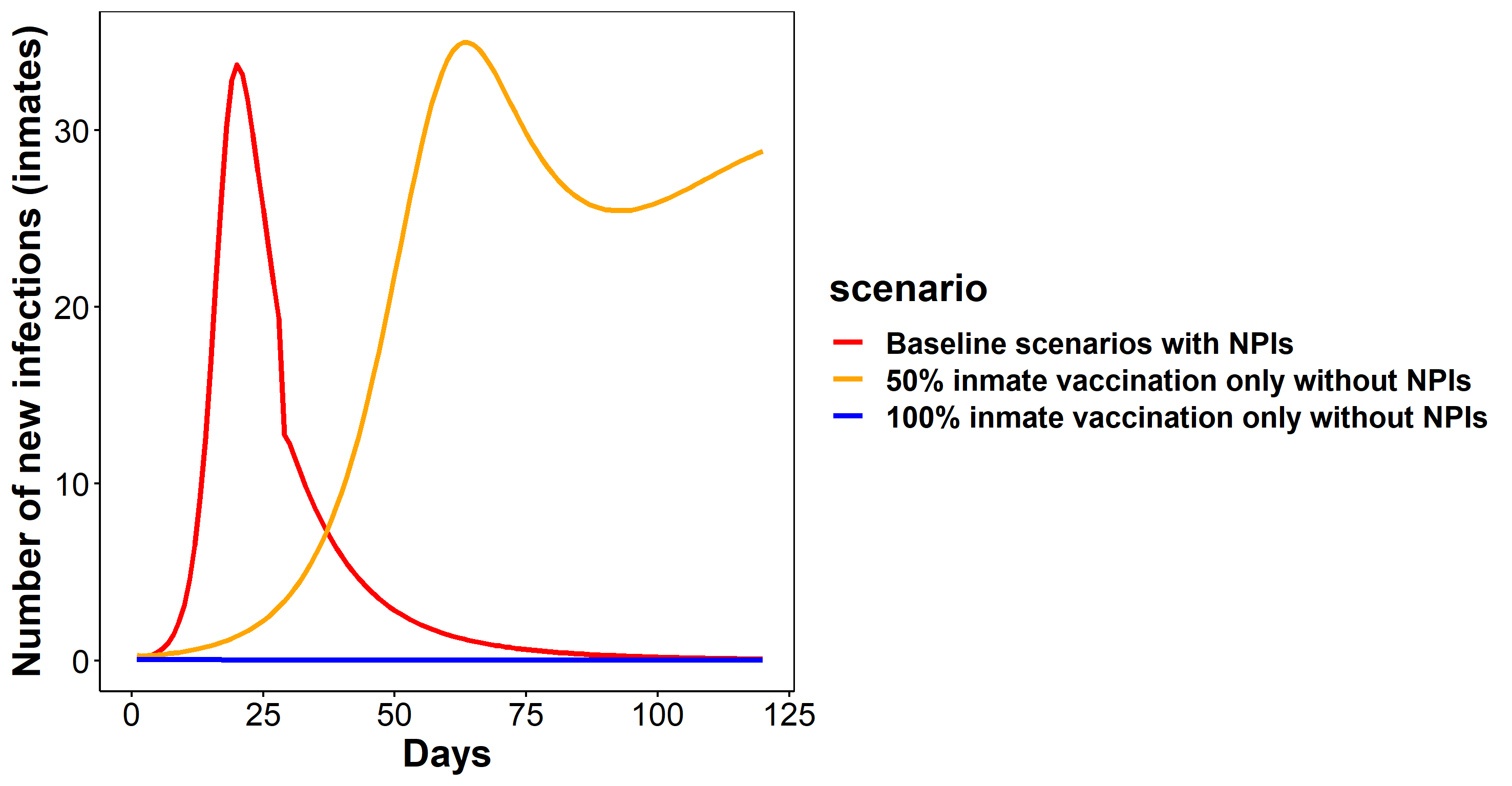 |
